# Clubfoot Twin Study with Wearable GaitUp Sensors and Footscan V9

**DOI:** 10.1101/2023.06.24.23291809

**Authors:** Issler-Wüthrich Ursula, Issler Christian, Exner G. Ulrich, Joller Peter

**Author notes:** Corresponding Author: Peter Joller, Spitzackerstrasse 8, 8057 Zürich, Switzerland.

## Abstract

**Purpose:** To control the therapeutic results in manual-dynamic physiotherapy for clubfeet we analyzed the gait pattern in children with clubfeet and their healthy twin siblings, aged between 3 and 13 years for GaitUp and 4 to 14 years for the footplate V9.

**Methods:** With the inertial GaitUp sensors and the footscan V9 pedobarographic plate the 11 twin-pairs were tested and statistically assessed. For the GaitUp sensors 22 parameters were considered and 10 parameters for the footplate V9. We analyzed the gait pattern for each child separately for both feet and in a second evaluation compared the affected feet with the ipsilateral feet of the healthy twins. The statistical comparisons were made with nonparametric methods. An additional twin girl treated with various therapies and her sister are included as a contrast.

**Results:** Especially in younger children, the gait pattern is not stabilized yet. Therefore, sometimes the healthy twins have inferior values in gait patterns than the affected siblings. Over the whole study there are only minor statistical differences between the affected group and the healthy group suggesting that with the manual-dynamic therapy the clubfeet children show a gait pattern statistically similar to the healthy group. Noteworthy are the less convincing results of the contrast twin.

**Conclusion:** Manual-dynamic physiotherapy can lead to a gait pattern equal to the one of unaffected children the same age. Our results do not support the statement that in one-sided clubfoot the other foot cannot be considered normal.

**IMPLICATIONS FOR REHABILITATION:**

- Clubfeet are a disabling birth defect affecting 1 to 3 babies per 1000 births.
- The manual-dynamic physiotherapy for clubfeet starting at birth leads to a normal gait pattern.

## Introduction

Clubfoot is the most common congenital deformity of the lower extremity with a number of permutations and combinations and already described by Hippocrates 400 BC [1]. The foot deformities interfere severely with locomotion ability and gait pattern and can lead to lifelong disability if not treated adequately. Human gait is an aspect with extraordinary complexity. Individual movements occurring simultaneously in the three planes of space make analysis difficult [2]. Quite a few publications stress the importance of computerized gait analysis in clubfeet for quality control [3,4,5,6]. Just visual observation of the walking pattern is too subjective [7]. Gait analysis in specialized laboratories has the disadvantage of being a foreign surrounding for the children and therefore influences the way of walking [8]. Computerized, wearable sensors and corresponding algorithms enable three dimensional spatio-temporal gait analysis and allow precise measurements in a doctor’s office with minimal effort [9]. Another aspect of the evaluation of gait patterns is the plantar pressure assessment of human walking. Modern pedobarographic plates and adequate software make quantitative interpretation of loading of the foot available for doctor’s offices [10].

In our practice with the footscan V9 (kinetics) and the wearable inertial GaitUp sensors (kinematics) we control the therapeutic results of the clubfoot treatment. Digital-quantitative methods are the only way to describe subtle deviations from normal indicating a pending relapse even after 5 years of age [11].

In the literature it is described that in one-sided clubfoot, the contralateral foot cannot be considered as comparable to a normal foot [12,13]. To test this statement, we selected for gait analysis eleven twin pairs from our patient collective where one sibling is affected, the other one is healthy.

## MATERIAL AND METHODS

### Ethics

This study has been approved by the Ethics committee of Zurich, Switzerland, Number 2023-00290. ClinicalTrials ID: NCT05913934.

### Patients

The parents of 11 pairs of twins and the contrast pair gave written informed consent for the participation for these control visits with inertial sensors and the footscan. All children involved gave their oral consent. The patients (15 clubfeet) of our group have been treated to date with manual-dynamic physiotherapy (Manuelles Zürcher Klumpfuss Konzept) by IU [14]. For comparison the analyses of a twin pair were one girl has double sided clubfeet treated elsewhere is included.

**Table 1.**
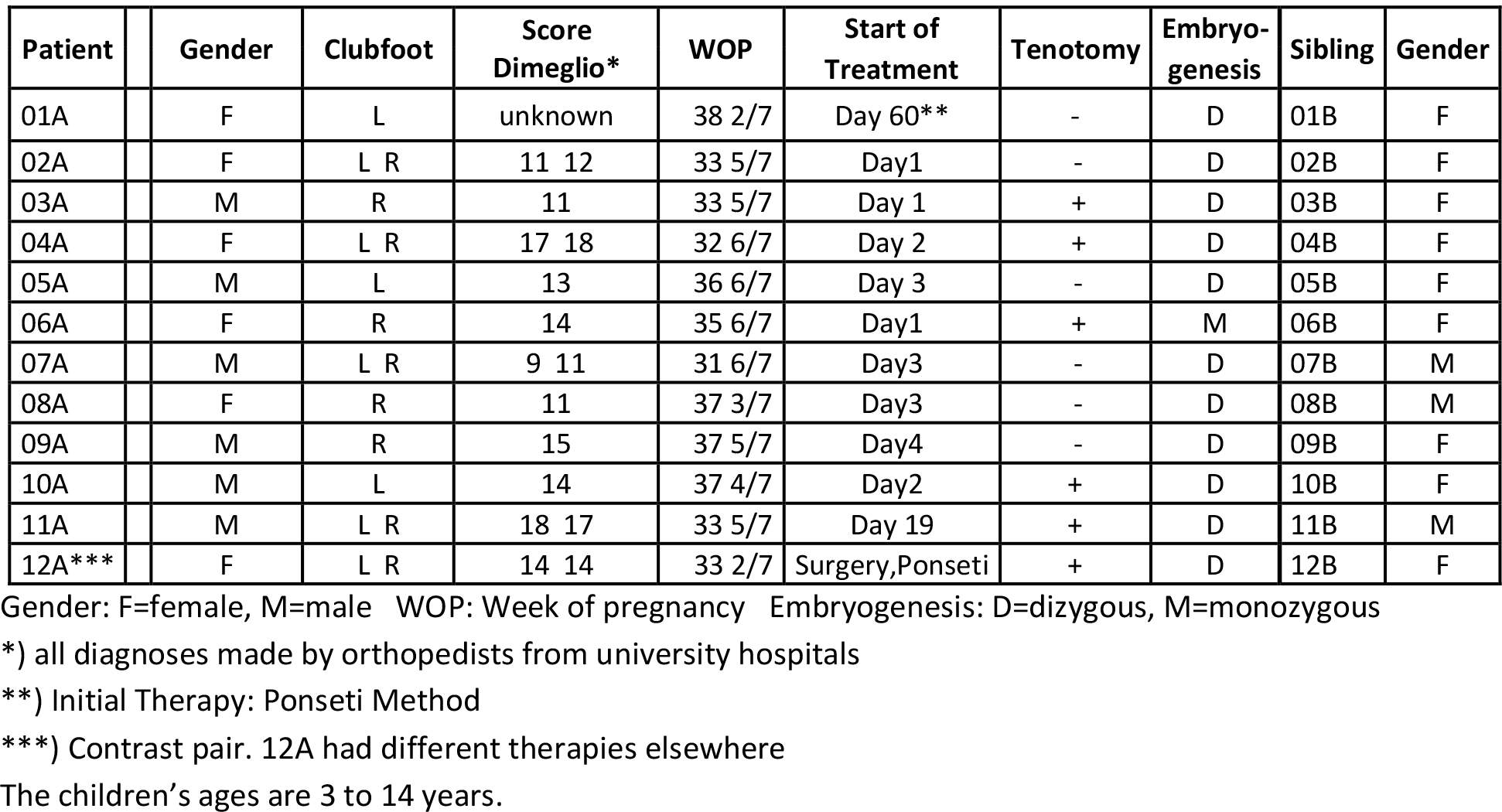
Anthropometric details.

### Gait Analysis Systems

**Fig. 1.**
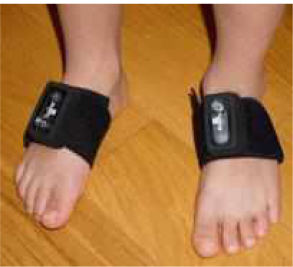
GaitUp sensors.

GaitUp sensors are wireless, inertial sensors (11g, self-calibrating) with a 3D accelerometer and a 3D gyroscope with 10D sensing capabilities (GaitUp, EPFL Innovation Park Bâtiment C, CH-1015 Lausanne, Switzerland). The sensors can yield 22 kinematic gait parameters. The sensors are attached to each foot with Velcro bands and are automatically synchronized. They are well accepted by children two years of age or older. GaitUp sensors have been validated with a motion capture system with 7 cameras (Vicon, UK) [15]. Most applications of GaitUp sensors until now were in adults. In children cerebral palsy was assessed [16].

In our study the test for each child comprises a walk of 25m in a gymnasium, repeated 3 times on a flat, even surface at a self-selected speed. First test series with shoes, second test series barefoot.

**Table 2.**
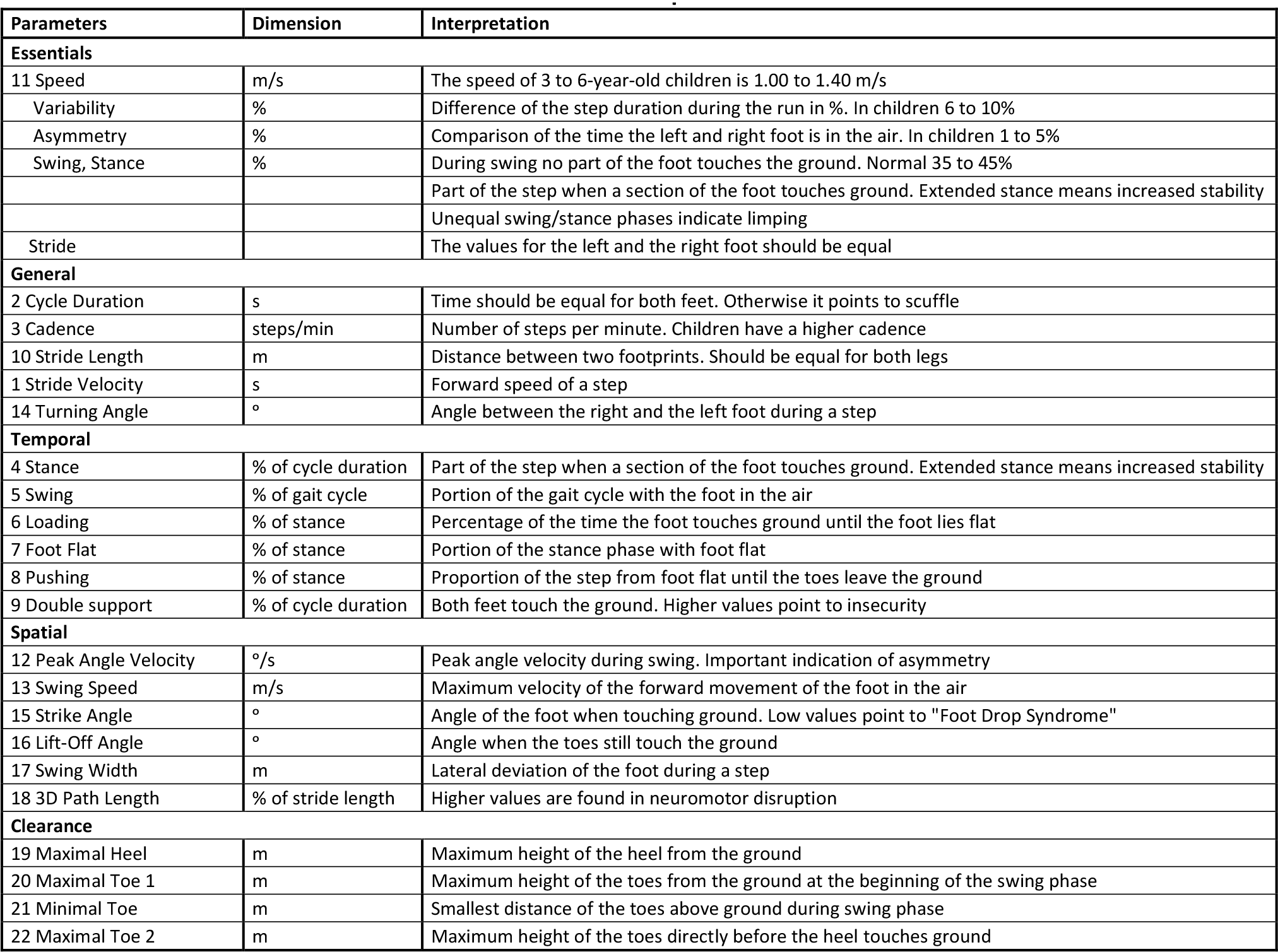
Measurement values of the GaitUp Sensors and their interpretation.

**Fig. 2.**
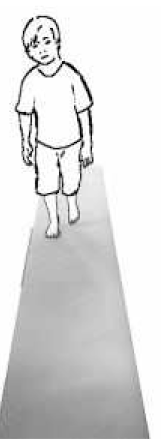
Footscan V9.

Plantar pressure plate: 4096 sensors; scanning rate 300 measurements per second. (RSscan Lab Ltd.10-15 Pegasus, Orion Court, Great Blakenham Suffolk, England).

This pressure plate is widely accepted and reliable [17]. We used a top-layer of ethylene-vinyl acetate copolymer (EVA) material (hardness: ShoreA 70, 2mm) on the plate and on the path. This layer hides the position of the pressure plate and circumvents the children trying to strike the plate with their feet which will result in an unnatural gait.

The walk is inspected on the computer screen and the trial is terminated when sufficient steps in good quality have been acquired.

To assess the differences between the clubfoot patients and the healthy sibling we selected for our study the following footscan V9 parameters:

Exorotation, Minimum Subtalar Joint Angle, Maximum Subtalar Joint Angle, Subtalar Joint Flexibility, Foot Length and Foot Width, Initial Contact Phase, Forefoot Contact Phase, Foot Flat Phase and Forefoot Push Off Phase. For the comparisons with 12A and 12B we analyzed the forces curves of M2, M3, MH and LH.

**Table 3.** Measurement values of the footscan V9 and t<u>heir interpretation</u>.

|  | Dimension | Interpretation |
| --- | --- | --- |
| Exorotation | ° | Midline axis of the hindfoot and the forefoot. Deviation from 0° |
| Minimum Subtalar Joint Angle | ° | Describes supination or pronation |
| Maximum Subtalar Joint Angle | ° | Describes supination or pronation |
| Subtalar Joint Flexibility | ° | normal 39° to 49° (range: 36°- 61°) |
| Foot Length | cm | Both feet should have the same length |
| Foot Width | cm | Both feet should have the same width |
| Initial Contact Phase | ms / % | First 3% of the gait cycle. Heel touches ground until first metatarsal contact |
| Forefoot Contact Phase | ms / % | 14% of the gait cycle. From first metatarsal contact to when all metatarsal zones make contact |
| Foot Flat Phase | ms / % | 30% of the gait cycle. Heel and forefoot on the ground |
| Forefoot Push Off Phase | ms / % | 53% of the gait cycle. Heel off the ground, foot off plate |

**Fig. 3.**
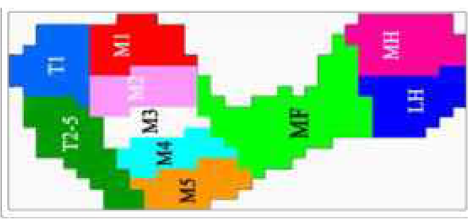
Foot zones for footscan V9. Our measurements for the comparison (M2, M3, MH and LH) are based on the work of Xu et al. [18]

### Statistics

Working hypothesis: With the digital-quantitative computer data obtained with the GaitUp sensors and the footscan V9 it will be possible to analyze the differences in the gait parameters between the group of healthy twins versus the group of the affected and identically treated siblings. Any significant disparity will be an indication to intensify or improve the therapy for the corresponding parameters. A second comparison was performed between the affected foot and the ipsilateral foot of the healthy twin siblings.

In this study we deal with a limited sample, therefore nonparametric statistical tools were used. For comparisons we worked with Wilcoxon signed rank test with Tukey’s post hoc procedure. Significance level α is 5%.

The concept of the effect size helps in the interpretation of the results. Due to the limited data we used Hedge’s g as a measure of the effect size. The effect size represents the shift of the two Gaussian curves compared in standard deviation units. A Hedge’s g of 0.2 is considered a small effect, 0.5 is a medium effect and 0.8 is a large effect.

Hedge’s g can also be negative when the shift is in the opposite direction.

To illustrate the disparity of the differently treated double-sided clubfoot twin we compared her results with her sister’s and with twins of a similar age and double-sided clubfeet of our patient group. Parameters are the footscan V9 forces curves of M2, M3, MH and LH.

Statistical calculations were done with Analyse-it (Analyse-it Software, Ltd., The Tannery, 91 Kirkstall Road, Leeds, LS3 1HS, United Kingdom).

## RESULTS

**Table 4.**
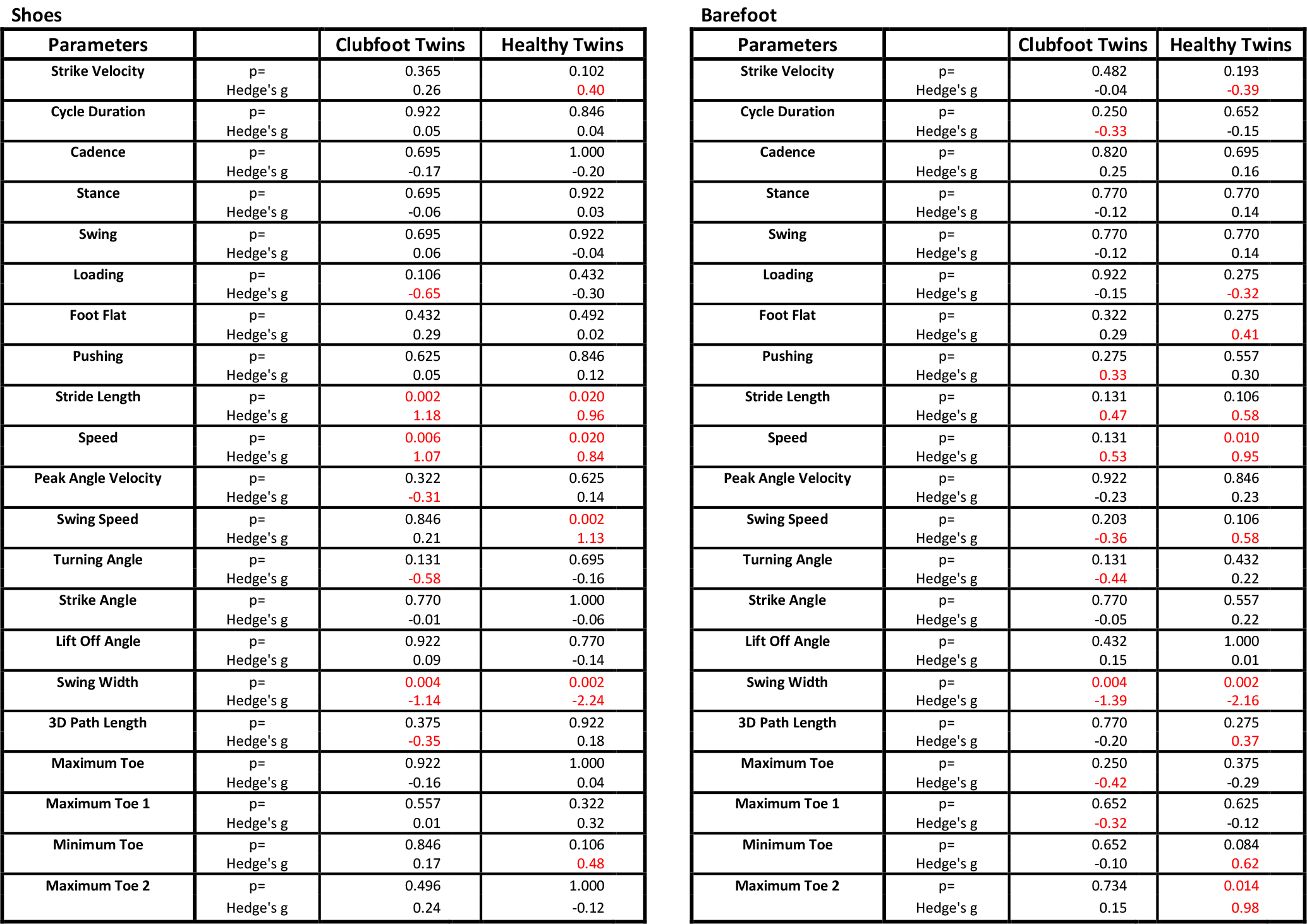
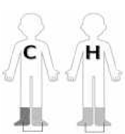 Comparison of the GaitUp parameters for both feet of the one-sided affected children (C) and the healthy twins (H) Single parameters quantified by the GaitUp sensors are statistically analyzed for the runs with shoes and barefoot. The comparison of both feet of the one-sided affected siblings and their healthy twins are assessed by the Wilcoxon signed rank test and the statistical Hedge’s g effect size. Wilcoxon p values below 0.05 point to a marked difference between left and right foot. Hedge’s g in excess of ±0.3 indicate a dissimilarity effect. The corresponding values are printed in red. Double support is not included because it is identical for both feet. (Interpretation of the parameters see Table 2).

| Shoes |  |  |  | Barefoot |  |  |  |
| --- | --- | --- | --- | --- | --- | --- | --- |
| Parameters |  | Clubfoot Twins | Healthy Twins | Parameters |  | Clubfoot Twins | Healthy Twins |
| Strike Velocity | p=<br>Hedge's $g$ | 0.365<br>0.26 | 0.102<br>**0.40** | Strike Velocity | p=<br>Hedge's $g$ | 0.482<br>-0.04 | 0.193<br>**-0.39** |
| Cycle Duration | p=<br>Hedge's $g$ | 0.922<br>0.05 | 0.846<br>0.04 | Cycle Duration | p=<br>Hedge's $g$ | 0.250<br>**-0.33** | 0.652<br>-0.15 |
| Cadence | p=<br>Hedge's $g$ | 0.695<br>-0.17 | 1.000<br>-0.20 | Cadence | p=<br>Hedge's $g$ | 0.820<br>0.25 | 0.695<br>0.16 |
| Stance | p=<br>Hedge's $g$ | 0.695<br>-0.06 | 0.922<br>0.03 | Stance | p=<br>Hedge's $g$ | 0.770<br>-0.12 | 0.770<br>0.14 |
| Swing | p=<br>Hedge's $g$ | 0.695<br>0.06 | 0.922<br>-0.04 | Swing | p=<br>Hedge's $g$ | 0.770<br>-0.12 | 0.770<br>0.14 |
| Loading | p=<br>Hedge's $g$ | 0.106<br>**-0.65** | 0.432<br>-0.30 | Loading | p=<br>Hedge's $g$ | 0.922<br>-0.15 | 0.275<br>**-0.32** |
| Foot Flat | p=<br>Hedge's $g$ | 0.432<br>0.29 | 0.492<br>0.02 | Foot Flat | p=<br>Hedge's $g$ | 0.322<br>0.29 | 0.275<br>**0.41** |
| Pushing | p=<br>Hedge's $g$ | 0.625<br>0.05 | 0.846<br>0.12 | Pushing | p=<br>Hedge's $g$ | 0.275<br>**0.33** | 0.557<br>0.30 |
| Stride Length | p=<br>Hedge's $g$ | **0.002**<br>**1.18** | **0.020**<br>**0.96** | Stride Length | p=<br>Hedge's $g$ | 0.131<br>**0.47** | 0.106<br>**0.58** |
| Speed | p=<br>Hedge's $g$ | **0.006**<br>**1.07** | **0.020**<br>**0.84** | Speed | p=<br>Hedge's $g$ | 0.131<br>**0.53** | **0.010**<br>**0.95** |
| Peak Angle Velocity | p=<br>Hedge's $g$ | 0.322<br>**-0.31** | 0.625<br>0.14 | Peak Angle Velocity | p=<br>Hedge's $g$ | 0.922<br>-0.23 | 0.846<br>0.23 |
| Swing Speed | p=<br>Hedge's $g$ | 0.846<br>0.21 | **0.002**<br>**1.13** | Swing Speed | p=<br>Hedge's $g$ | 0.203<br>**-0.36** | 0.106<br>**0.58** |
| Turning Angle | p=<br>Hedge's $g$ | 0.131<br>**-0.58** | 0.695<br>-0.16 | Turning Angle | p=<br>Hedge's $g$ | 0.131<br>**-0.44** | 0.432<br>0.22 |
| Strike Angle | p=<br>Hedge's $g$ | 0.770<br>-0.01 | 1.000<br>-0.06 | Strike Angle | p=<br>Hedge's $g$ | 0.770<br>-0.05 | 0.557<br>0.22 |
| Lift Off Angle | p=<br>Hedge's $g$ | 0.922<br>0.09 | 0.770<br>-0.14 | Lift Off Angle | p=<br>Hedge's $g$ | 0.432<br>0.15 | 1.000<br>0.01 |
| Swing Width | p=<br>Hedge's $g$ | **0.004**<br>**-1.14** | **0.002**<br>**-2.24** | Swing Width | p=<br>Hedge's $g$ | **0.004**<br>**-1.39** | **0.002**<br>**-2.16** |
| 3D Path Length | p=<br>Hedge's $g$ | 0.375<br>**-0.35** | 0.922<br>0.18 | 3D Path Length | p=<br>Hedge's $g$ | 0.770<br>-0.20 | 0.275<br>**0.37** |
| Maximum Toe | p=<br>Hedge's $g$ | 0.922<br>-0.16 | 1.000<br>0.04 | Maximum Toe | p=<br>Hedge's $g$ | 0.250<br>**-0.42** | 0.375<br>-0.29 |
| Maximum Toe 1 | p=<br>Hedge's $g$ | 0.557<br>0.01 | 0.322<br>0.32 | Maximum Toe 1 | p=<br>Hedge's $g$ | 0.652<br>**-0.32** | 0.625<br>-0.12 |
| Minimum Toe | p=<br>Hedge's $g$ | 0.846<br>0.17 | 0.106<br>**0.48** | Minimum Toe | p=<br>Hedge's $g$ | 0.652<br>-0.10 | 0.084<br>**0.62** |
| Maximum Toe 2 | p=<br>Hedge's $g$ | 0.496<br>0.24 | 1.000<br>-0.12 | Maximum Toe 2 | p=<br>Hedge's $g$ | 0.734<br>0.15 | **0.014**<br>**0.98** |

**Table 5.**
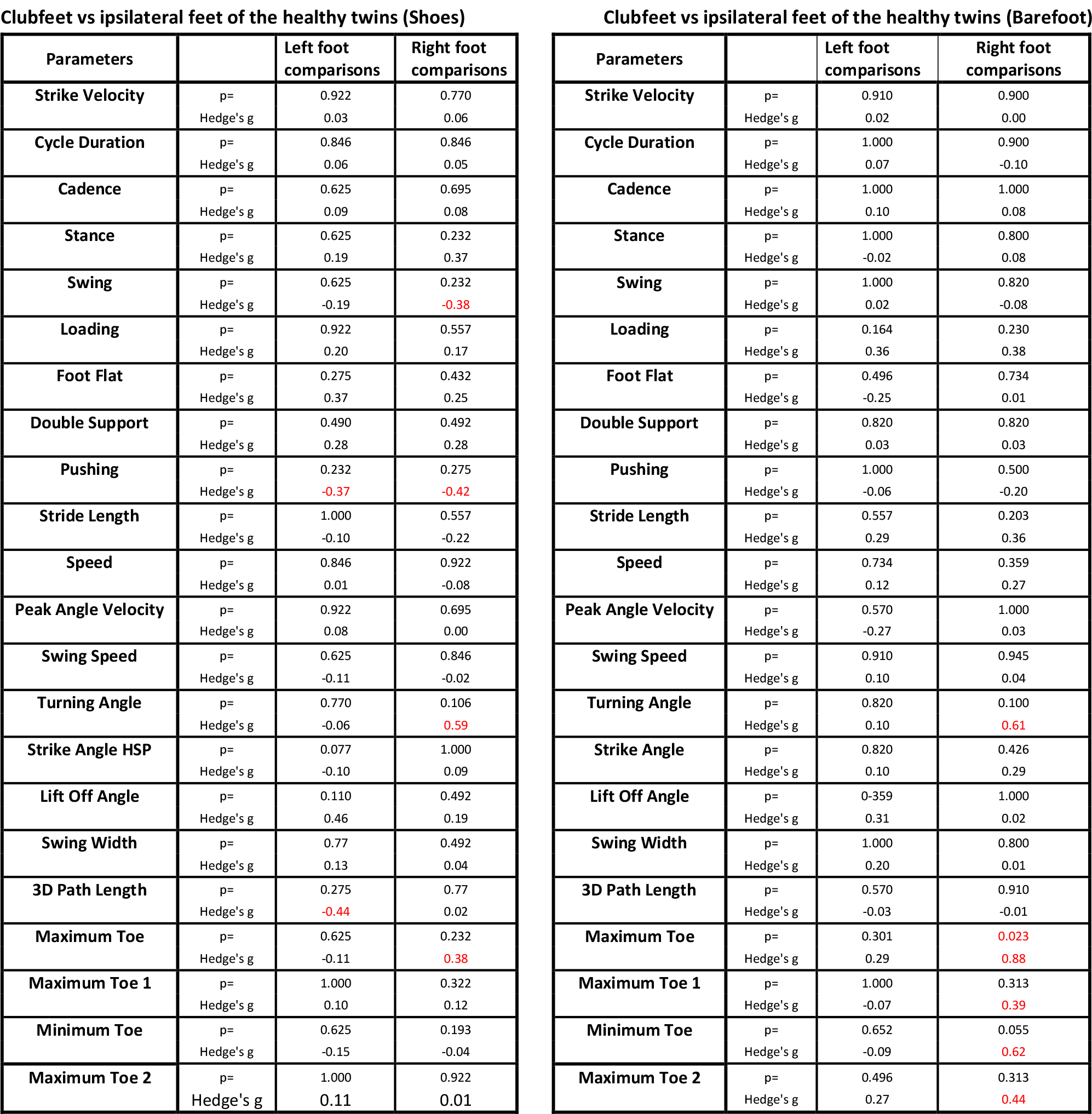
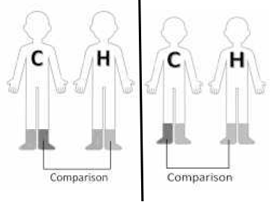 Comparison of the GaitUp parameters of the clubfoot of the affected twins (C) and the ipsilateral foot of the siblings (H). For the left feet there are 7 pairs, for the right feet 8 pairs. Comparison between all clubfeet with the ipsilateral feet of the healthy siblings in relation to the different parameters. (Interpretation of the parameters see Table 2).

**Clubfeet vs ipsilateral feet of the healthy twins (Shoes)**
| Parameters |  | Left foot comparisons | Right foot comparisons |
| --- | --- | --- | --- |
| Strike Velocity | p=<br>Hedge's g | 0.922<br>0.03 | 0.770<br>0.06 |
| Cycle Duration | p=<br>Hedge's g | 0.846<br>0.06 | 0.846<br>0.05 |
| Cadence | p=<br>Hedge's g | 0.625<br>0.09 | 0.695<br>0.08 |
| Stance | p=<br>Hedge's g | 0.625<br>0.19 | 0.232<br>0.37 |
| Swing | p=<br>Hedge's g | 0.625<br>-0.19 | 0.232<br>**-0.38** |
| Loading | p=<br>Hedge's g | 0.922<br>0.20 | 0.557<br>0.17 |
| Foot Flat | p=<br>Hedge's g | 0.275<br>0.37 | 0.432<br>0.25 |
| Double Support | p=<br>Hedge's g | 0.490<br>0.28 | 0.492<br>0.28 |
| Pushing | p=<br>Hedge's g | 0.232<br>**-0.37** | 0.275<br>**-0.42** |
| Stride Length | p=<br>Hedge's g | 1.000<br>-0.10 | 0.557<br>-0.22 |
| Speed | p=<br>Hedge's g | 0.846<br>0.01 | 0.922<br>-0.08 |
| Peak Angle Velocity | p=<br>Hedge's g | 0.922<br>0.08 | 0.695<br>0.00 |
| Swing Speed | p=<br>Hedge's g | 0.625<br>-0.11 | 0.846<br>-0.02 |
| Turning Angle | p=<br>Hedge's g | 0.770<br>-0.06 | 0.106<br>**0.59** |
| Strike Angle HSP | p=<br>Hedge's g | 0.077<br>-0.10 | 1.000<br>0.09 |
| Lift Off Angle | p=<br>Hedge's g | 0.110<br>0.46 | 0.492<br>0.19 |
| Swing Width | p=<br>Hedge's g | 0.77<br>0.13 | 0.492<br>0.04 |
| 3D Path Length | p=<br>Hedge's g | 0.275<br>**-0.44** | 0.77<br>0.02 |
| Maximum Toe | p=<br>Hedge's g | 0.625<br>-0.11 | 0.232<br>**0.38** |
| Maximum Toe 1 | p=<br>Hedge's g | 1.000<br>0.10 | 0.322<br>0.12 |
| Minimum Toe | p=<br>Hedge's g | 0.625<br>-0.15 | 0.193<br>-0.04 |
| Maximum Toe 2 | p=<br>Hedge's g | 1.000<br>0.11 | 0.922<br>0.01 |

Table 5. Comparison between all clubfeet with the ipsilateral feet of the healthy siblings in relation to the different parameters. (Interpretation of the parameters see Table 2).
| Parameters |  | Left foot comparisons | Right foot comparisons |
| --- | --- | --- | --- |
| Strike Velocity | p=<br>Hedge's g | 0.910<br>0.02 | 0.900<br>0.00 |
| Cycle Duration | p=<br>Hedge's g | 1.000<br>0.07 | 0.900<br>-0.10 |
| Cadence | p=<br>Hedge's g | 1.000<br>0.10 | 1.000<br>0.08 |
| Stance | p=<br>Hedge's g | 1.000<br>-0.02 | 0.800<br>0.08 |
| Swing | p=<br>Hedge's g | 1.000<br>0.02 | 0.820<br>-0.08 |
| Loading | p=<br>Hedge's g | 0.164<br>0.36 | 0.230<br>0.38 |
| Foot Flat | p=<br>Hedge's g | 0.496<br>-0.25 | 0.734<br>0.01 |
| Double Support | p=<br>Hedge's g | 0.820<br>0.03 | 0.820<br>0.03 |
| Pushing | p=<br>Hedge's g | 1.000<br>-0.06 | 0.500<br>-0.20 |
| Stride Length | p=<br>Hedge's g | 0.557<br>0.29 | 0.203<br>0.36 |
| Speed | p=<br>Hedge's g | 0.734<br>0.12 | 0.359<br>0.27 |
| Peak Angle Velocity | p=<br>Hedge's g | 0.570<br>-0.27 | 1.000<br>0.03 |
| Swing Speed | p=<br>Hedge's g | 0.910<br>0.10 | 0.945<br>0.04 |
| Turning Angle | p=<br>Hedge's g | 0.820<br>0.10 | 0.100<br>**0.61** |
| Strike Angle | p=<br>Hedge's g | 0.820<br>0.10 | 0.426<br>0.29 |
| Lift Off Angle | p=<br>Hedge's g | 0-359<br>0.31 | 1.000<br>0.02 |
| Swing Width | p=<br>Hedge's g | 1.000<br>0.20 | 0.800<br>0.01 |
| 3D Path Length | p=<br>Hedge's g | 0.570<br>-0.03 | 0.910<br>-0.01 |
| Maximum Toe | p=<br>Hedge's g | 0.301<br>0.29 | **0.023**<br>**0.88** |
| Maximum Toe 1 | p=<br>Hedge's g | 1.000<br>-0.07 | 0.313<br>**0.39** |
| Minimum Toe | p=<br>Hedge's g | 0.652<br>-0.09 | 0.055<br>**0.62** |
| Maximum Toe 2 | p=<br>Hedge's g | 0.496<br>0.27 | 0.313<br>**0.44** |

**Table 6.** 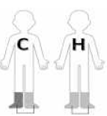 Comparison of the footscan V9 parameters for both feet of the one-sided affected children (C) and the healthy twins (H) The gait analysis of the footscan V9 parameters measured in the clubfeet children and their healthy siblings. Barefoot only. Wilcoxon p ≤ 0.05 describes a statistically significant difference for the respective parameter and are printed in red. Hedge’s g is the statistical effect size. g = ±0.2 is considered a small effect size, ±0.5 is a medium effect size and ±0.8 a large effect size. Values in excess of ±0.3 are printed in red. (Interpretation of the parameters see Table 3).

| Parameters |  | Clubfoot Twins | Healthy Twins |
| --- | --- | --- | --- |
| Exorotation | p= | 0.913 | 0.413 |
|  | Hedge's g | -0.14 | -0.22 |
| Minimum subtalar joint angle | p= | 0.765 | 1.000 |
|  | Hedge's g | 0.10 | 0.00 |
| Maximum subtalar joint angle | p= | 0.496 | 0.375 |
|  | Hedge's g | -0.10 | -0.10 |
| Subtalar joint flexibility | p= | 0.625 | 0.820 |
|  | Hedge's g | -0.20 | 0.20 |
| Foot Length | p= | 0.250 | 0.734 |
|  | Hedge's g | -0.23 | 0.08 |
| Foot Width | p= | 0.922 | 0.922 |
|  | Hedge's g | 0.04 | 0.03 |
| Initial Contact Phase | p= | 0.813 | 0.193 |
|  | Hedge's g | 0.00 | 0.40 |
| Forefoot Contact Phase | p= | 0.570 | 0.426 |
|  | Hedge's g | -0.30 | -0.30 |
| Foot Flat Phase | p= | 0.106 | 0.123 |
|  | Hedge's g | 0.60 | -0.40 |
| Forefoot Push Off Phase | p= | 0.193 | 0.067 |
|  | Hedge's g | -0.40 | 0.60 |

**Table 7.** 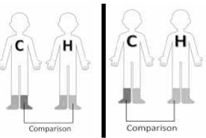 Comparison of single footscan V9 parameters between the affected feet and the ipsilateral feet of the healthy twins. For the left feet there are 7 pairs, for the right feet 8 pairs. Comparison of the affected feet of the siblings with the ipsilateral feet of the healthy twins.

| Parameters |  | Left foot comparisons | Right foot comparisons |
| --- | --- | --- | --- |
| Exorotation | p= | 0.467 | 0.313 |
|  | Hedge's g | 0.62 | -0.50 |
| Minimum subtalar joint angle | p= | 0.844 | 1.000 |
|  | Hedge's g | 0.10 | -0.10 |
| Maximum subtalar joint angle | p= | 0.109 | 0.461 |
|  | Hedge's g | 0.60 | 0.30 |
| Subtalar joint flexibility | p= | 0.438 | 0.461 |
|  | Hedge's g | 0.40 | 0.30 |
| Foot Length | p= | 0.688 | 0.547 |
|  | Hedge's g | -0.17 | 0.22 |
| Foot Width | p= | 0.938 | 0.578 |
|  | Hedge's g | -0.21 | 0.03 |
| Initial Contact Phase | p= | 0.094 | 0.250 |
|  | Hedge's g | 0.80 | 0.50 |
| Forefoot Contact Phase | p= | 0.563 | 0.055 |
|  | Hedge's g | -0.20 | 0.70 |
| Foot Flat Phase | p= | 0.313 | 0.813 |
|  | Hedge's g | 0.40 | 0.00 |
| Forefoot Push Off Phase | p= | 0.219 | 0.945 |
|  | Hedge's g | -0.40 | -0.10 |

**Fig. 4.**
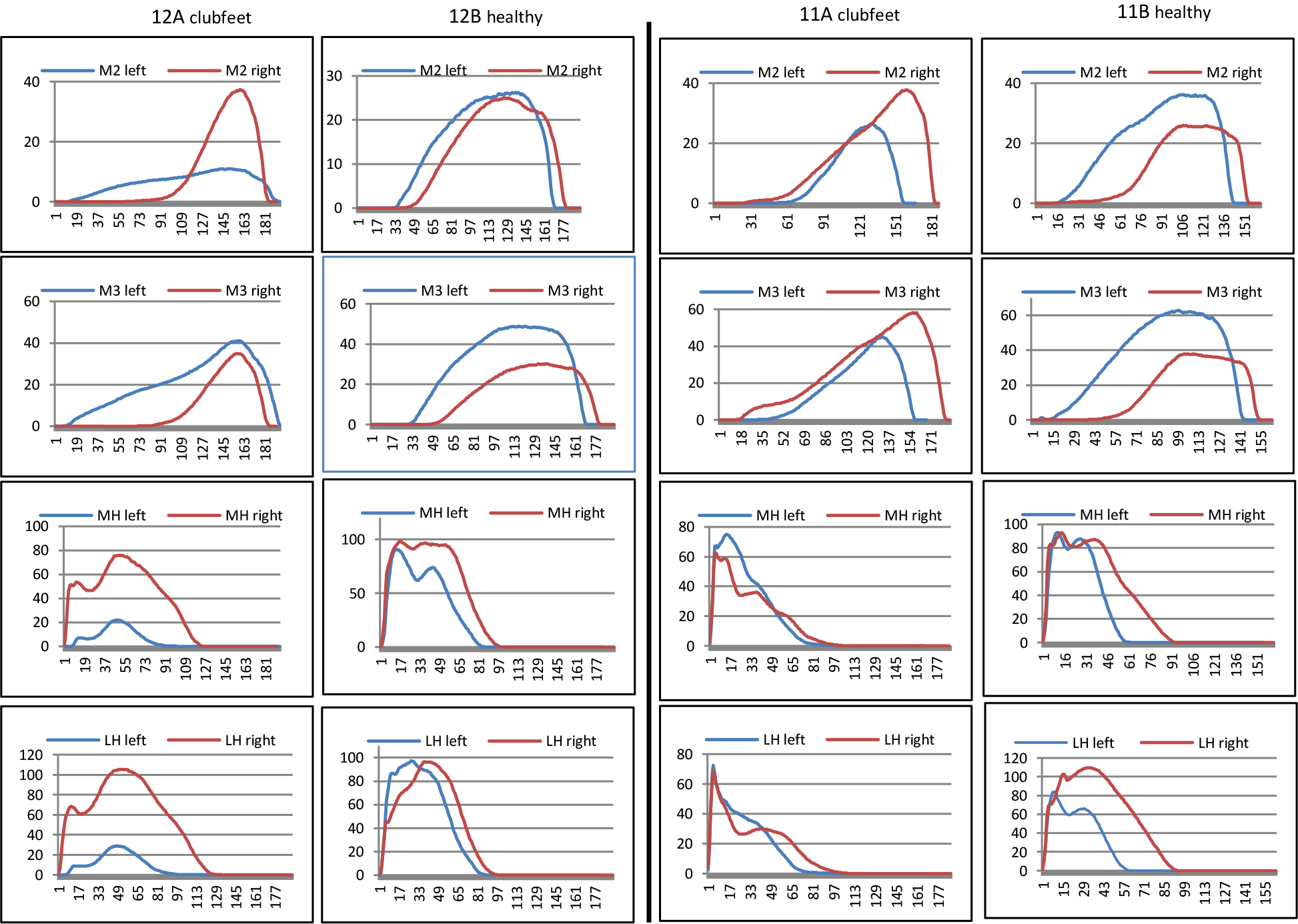
Comparison: Child with inadequate therapy (12A), her sister (12B) and a twin pair of our group (11A; 11B) From the footscan V9 data the forces values for M2, M3 MH and LH were extracted and depicted as curves for the left and the right foot. The double-sided clubfeet of (12A) were not treated by UI with the manual-dynamic therapy. 12A is compared with her sister (12B) and one of our pairs the same age and double-sided.

## Discussion

To our knowledge this is the first comparative study with a cohort of clubfoot twins considering gait analysis with two independent systems following identical treatment by a single person. Other twin publications concentrate on male to female ratio (2.5 to 1), twinning effects (2.9% versus 1.2% in the population) and discrepancy between monozygotic versus dizygotic twins (less clubfeet in dizygotic twins)[19].

Our working hypothesis for the gait analysis was finding parameters not considered normal to adapt the treatment accordingly. However, we couldn’t find consistent deviations between the clubfoot children and their healthy twins. The reason might be the limited sample. Among 300 clubfeet we had 11 twin-pairs for investigation. The few statistically significant gait differences might also be due to the varying ages from 3 to 14 years. If in these instances we split the youngest 5 pairs from the 6 older pairs the statistical significance disappears (data not shown). The majority of the gait pattern parameters analyzed in our patient group show an equivalence between the clubfoot children treated with manual-dynamic physiotherapy and their healthy twins. One goal of clubfoot therapy is to reach the physiologic derotation as measured with the footscan V9 exorotation. This parameter shows no statistical significant difference between the affected and the healthy children.

The clubfeet of a girl treated with varying methods reveals a substantial difference to the manual-dynamic treated group. The deviations in this girl support the validity of the gait analyses with the pedobarometric plate. Unfortunately, normal values for children in the field of gait patterns with similar methods are missing. Such data would be beneficial to further validate the presented data.

Today there are different accepted modalities to treat clubfeet. An important principle of the manual-dynamic method is to avoid thigh-casts and immobilizing (night-) braces [14]. We assume that this reduces negative effects on the child’s perception and development. The early beginning of the therapy may reduce fibrosis of connective tissues due to impaired movement during pregnancy. The derotating lower leg orthosis used allow for the knee- and hip mobility. [14]. The three-dimensional torsion of the clubfoot is multi-dimensionally well addressed with the manual-dynamic therapy in all levels and axes. Thus, the calcaneotalar angle in the transverse plane could be adjusted to normal values. Highly important is the inclusion of the functional movement chain in the treatment.

Admittedly our study is a snapshot and permits no statement for a lifelong absence of symptoms, the ultimate goal of every therapy. There is a generally accepted need for further studies.

The key of our study design lays in the fact that the controls and patients share the exact same age, the same environment and genetics. This ensures statistical homogeneity. The drawback is the small sample size. Future studies are needed.

## Data Availability

All data are with the authors and are available anonymized upon reasonable request

## References

[1] A. Anand and D. Sala, “Clubfoot: Etiology and treatment,” Indian J. Orthop., 2009, doi: 10.4103/0019-5413.38576.

[2] C. P. Charalambous, “The major determinants in normal and pathological gait,” in Classic Papers in Orthopaedics, 2014, pp. 403–405.

[3] M. Sangeux, E. Passmore, H. K. Graham, and O. Tirosh, “The gait standard deviation, a single measure of kinematic variability,” Gait Posture, vol. 46, no. March, pp. 194–200, 2016, doi: 10.1016/j.gaitpost.2016.03.015.

[4] T. Alkjær, E. N. G. Pedersen, and E. B. Simonsen, “Evaluation of the walking pattern in clubfoot patients who received early intensive treatment,” J. Pediatr. Orthop., vol. 20, no. 5, pp. 642–647, 2000, doi: 10.1097/00004694-200009000-00018.

[5] L. A. Karol, K. Jeans, and R. Elhawary, “Gait analysis after initial nonoperative treatment for clubfeet: Intermediate term followup at age 5,” in Clinical Orthopaedics and Related Research, 2009, vol. 467, no. 5, pp. 1206–1213, doi: 10.1007/s11999-008-0702-9.

[6] Feng Jing, “Application of gait analysis in pediatric orthopedics,” Curr. Orthop. Pract., vol. 27, no. 4, pp. 455– 464, 2016.

[7] W. N. Sankar, S. A. Rethlefsen, J. Weiss, and R. M. Kay, “The recurrent clubfoot: Can gait analysis help us make better preoperative decisions?,” Clin. Orthop. Relat. Res., vol. 467, no. 5, pp. 1214–1222, 2009, doi: 10.1007/s11999-008-0665-x.

[8] A. Brégou Bourgeois, B. Mariani, K. Aminian, P. Y. Zambelli, and C. J. Newman, “Spatio-temporal gait analysis in children with cerebral palsy using, foot-worn inertial sensors,” Gait Posture, vol. 39, no. 1, pp. 436–442, 2014, doi: 10.1016/j.gaitpost.2013.08.029.

[9] W. Tao, T. Liu, R. Zheng, and H. Feng, “Gait analysis using wearable sensors,” Sensors, vol. 12, no. 2. pp. 2255– 2283, 2012, doi: 10.3390/s120202255.

[10] K. Yu and F. Kazuo, “Quantitative analysis of plantar load distribution with comparison to ground reaction force during take-off phase in vertical jump movement,” Japanese J. Phys. Fit. Sport. Med., vol. 61, no. 3, pp. 351–363, 2012, doi: 10.7600/jspfsm.61.351.

[11] M. Bent et al., “Gait Analysis Characteristics in Relapsed Clubfoot,” J. Pediatr. Orthop., vol. 43, no. 2, pp. 65– 69, 2023, doi: 10.1097/BPO.0000000000002314.

[12] A. Cooper, H. Chhina, A. Howren, and C. Alvarez, “The contralateral foot in children with unilateral clubfoot, is the unaffected side normal?,” Gait Posture, vol. 40, no. 3, pp. 375–380, 2014, doi: 10.1016/j.gaitpost.2014.05.004.

[13] E. Lööf, H. Andriesse, M. André, S. Böhm, and E. W. Broström, “Gait in 5-year-old children with idiopathic clubfoot: A cohort study of 59 children, focusing on foot involvement and the contralateral foot,” Acta Orthop., vol. 87, no. 5, pp. 522–528, Sep. 2016, doi: 10.1080/17453674.2016.1202013.

[14] U. Issler, Klumpfusstherapie nach Issler-Wüthrich, 1st ed. In-house publisher, 2020. doi:10.5281/zenodo.7316562

[15] H. Schwameder, M. Andress, E. Graf, and G. Strutzenberger, “VALIDATION OF AN IMUNSYSTEM (GAIT-UP) TO IDENTIFY GAIT PARAMETERS IN NORMAL AND INDUCED LIMPING WALKING CONDITIONS.”

[16] L. Carcreff et al., “Comparison of gait characteristics between clinical and daily life settings in children with cerebral palsy,” Sci. Rep., vol. 10, no. 1, Dec. 2020, doi: 10.1038/s41598-020-59002-6.

[17] C. Xu et al., “Reliability of the Footscan® Platform System in Healthy Subjects: A Comparison of without Top-Layer and with Top-Layer Protocols,” Biomed Res. Int., vol. 2017, 2017, doi: 10.1155/2017/2708712.

[18] C. Xu et al., “Pedobarographic Analysis following Ponseti Treatment for Unilateral Neglected Congenital Clubfoot,” Sci. Rep., vol. 8, no. 1, Dec. 2018, doi: 10.1038/s41598-018-24737-w.

[19] C. Lochmiller, D. Johnston, A. Scott, M. Risman, and J. T. Hecht, “Genetic epidemiology study of idiopathic talipes equinovarus,” Am. J. Med. Genet., vol. 79, no. 2, pp. 90–96, 1998, doi: 10.1002/(SICI)1096-8628(19980901)79:2<90::AID-AJMG3>3.0.CO;2-R.

